# Evaluating eating behaviour, energy homeostasis and obesity in childhood onset craniopharyngioma: A feasibility study

**DOI:** 10.1101/2023.01.10.23284332

**Authors:** Elanor C. Hinton, Fiona E. Lithander, Rebecca L. Elsworth, Katherine Hawton, Kruthika Narayan, Sophie Szymkowiak, Holly L. Bedford, Nimra Naeem, Ruth Elson, Tashunka Taylor-Miller, Julian P. Hamilton-Shield, Elizabeth C. Crowne

## Abstract

**Introduction:** Craniopharyngiomas are rare brain tumours (incidence 1.1-1.7 cases/million/year). Although benign, craniopharyngioma causes major endocrine and visual morbidities including hypothalamic obesity, yet mechanisms leading to obesity are poorly understood. This study investigated the feasibility and acceptability of eating behaviour measures in patients with craniopharyngioma, to inform the design of future trials.

**Methods:** Patients with childhood-onset craniopharyngioma and controls matched for sex, pubertal stage and age were recruited. After an overnight fast, participants received the following measures: body composition, resting metabolic rate, oral-glucose-tolerance-test including MRI (patients only), appetite ratings, eating behaviour and quality of life questionnaires, ad libitum lunch, and an acceptability questionnaire. Data are reported as median ± IQR, with effect size measure (Cliff’s delta) and Kendall’s Tau for correlations, due to the small sample size.

**Results:** Eleven patients (median age=14yrs; 5F/6M) and matched controls (median age=12yrs; 5F/6M) were recruited. All patients had received surgery and 9/11 also received radiotherapy. Hypothalamic damage post-surgery was graded (Paris grading): grade 2 n=6; grade 1 n=1; grade 0 n=2. The included measures were deemed highly tolerable by participants and their parent/carers. Preliminary data suggests a difference in hyperphagia between patients and controls (d=0.5), and a relationship between hyperphagia with body mass index (BMISDS) in patients (τ=0.46).

**Discussion:** These findings demonstrate that eating behaviour research is feasible and acceptable to craniopharyngioma patients and there is an association between BMISDS and hyperphagia in patients. Thus, food approach and avoidance behaviours may be useful targets for interventions to manage obesity in this patient group.

## Introduction

Craniopharyngiomas are embryological, histologically benign brain tumours, located in the sellar and parasellar regions (1). The incidence is 0.5-2 new cases per million persons per year (2), of which 30-50% occur in children and adolescents (3). Survival is high, at over 98% (4), but with significant levels of morbidity. Craniopharyngioma and its treatments, including gross total resection, limited resection and/or irradiation (5), commonly lead to endocrine deficiencies, visual impairment and obesity (6).

Obesity is a common feature of childhood craniopharyngioma (50%), which may develop several years before diagnosis (7), though there is a marked increase in prevalence (22-62%) following surgery (8, 9). Obesity in childhood craniopharyngioma is associated with a high risk of multisystem morbidity (10, 11), metabolic syndrome (12), cardiovascular disease (13) and mortality (14). Yet, there is limited evidence for effective weight loss interventions (15) and pharmacological treatments (16, 17) in this population.

Mechanisms leading to obesity and altered energy homeostasis in craniopharngioma are complex, yet understudied. Hypothalamic involvement of the tumour or damage to the hypothalamus following treatment is associated with severe obesity (7) and the degree of obesity is positively correlated with extent of hypothalamic damage (18, 19). Evidence to date, suggests an association between overweight and obesity with hyperphagia (20-22), contrasting with other evidence that presents lower energy and fat intake compared to body mass index (BMI) matched controls (23). The extent of hypothalamic damage across patient groups (24) or methodological limitations could explain the discrepancy between these findings, including the number of patients willing and able to accurately complete self-reported dietary intake diaries (21). Alternative measures that could be employed to assess eating behaviour are the ad libitum meal, which has only been used by one research group with craniopharyngioma patients to date (17, 25), and validated psychological questionnaires that have been specifically designed to measure different eating behaviour traits (e.g. Hyperphagia (26); Child Eating Behaviour Questionnaire (27)). A scoping review exploring the current literature addressing eating behaviour and psychological mechanisms associated with obesity in patients with craniopharyngioma is currently in progress (28).

Beyond eating behaviour per se, evidence thus far suggests a reduction in energy expenditure following craniopharyngioma or its treatment (29-31), which may be mediated by damage to efferent sympathetic nervous system pathways in the hypothalamus (32). A preliminary study using functional neuroimaging suggested patients with craniopharyngioma may have altered food cue reactivity (25). Moreover, there is mixed evidence for differences in peripherally secreted hormones, such as leptin, GLP-1, insulin, PYY and ghrelin (19, 33-35). Inconsistencies in findings could be due to the differing extent of hypothalamic damage or levels of BMI-SDS in patient groups across studies.

Obesity in children and young people with craniopharyngioma causes substantial personal and parental worry (36). Quality of life (QoL) is negatively affected overall by craniopharyngioma (37), and obesity has been found to be a significant predictor of long-term QoL in paediatric patients (38). QoL as rated by the parents has been found to be more impaired compared to their child’s rating (36, 39). Therefore, investigating what factors may be associated with poor QoL in craniopharyngioma is an important focus of research to inform future interventions.

This feasibility study was designed to address four objectives to assess: (i) accessibility to medical records to fully characterise the patients’ eating behaviour and obesity to date, (ii) recruitment to research in this young patient group, and (iii) their tolerability and acceptability of a range of measures. By analysing the preliminary data from this study, including data from a control group matched on age, sex and pubertal stage, we also addressed the final objective (iv) to assess which measures might be the ‘most informative’ to characterise the eating behaviours of patients with childhood-onset craniopharyngioma to take forward into a future, multicentre trial. By ‘most informative’, we mean whether preliminary evidence was seen for a difference between those with and without craniopharyngioma, and/or a relationship with BMI SDS. This paper focuses on feasibility and acceptability of all the included measures, and preliminary data from the energy intake and expenditure measures. The larger dataset including neuroimaging and measures of peripheral entero-endocrine hormones results will be presented elsewhere. This study was registered on ISRCTN database (https://doi.org/10.1186/ISRCTN86005167).

## Materials and Methods

The study was conducted between January 2019 and October 2021. The protocol and associated documentation were approved by South West - Frenchay Research Ethics Committee (approval number 18/SW/0235; Nov 2018). When seeking informed consent, an age-appropriate patient information sheet was sent out to participants and parents/carers at least two weeks in advance of their next appointment, to allow time to consider participation. Potential participants and their parents were given the opportunity to ask any questions and the study was explained in a way the potential participant could understand. If participants aged 16 or over agreed to take part, they gave written informed consent on the form provided. Parents/carers of participants aged 16 or over were also informed about the study if the participants were happy for their parents/carers to be informed. If parents/carers were present at the appointment, they were also asked to give their consent (which was the case for the only 16+ participants in the study). Parents/carers were invited to provide written informed consent for those participants aged 15 or under, who provided assent to participate. Participants were free to withdraw at any time during the study without any impact on their medical care.

### Participants

Inclusion criteria: Craniopharyngioma diagnosed under 18 years of age, now currently aged 7-25 years. Exclusion criteria: clinically unwell requiring hospital or intensive treatment; unwilling to fast; significant visual impairment so unable to undertake food cue reactivity tasks; patients for whom it would be unsafe to have an MRI including those with metal implants, tattoos with metallic ink, or metal body piercings which cannot be removed; pregnancy to avoid harm to the fetus; claustrophobia in the closed MRI environment or unable to tolerate the MRI scanner; weight above 152kg and/or girth greater than 210cm due to size limitations of MRI scanner.

Following recruitment of the patient group, control participants were recruited matched on (i) sex (male/female at birth), (ii) pubertal stage (pre-pubertal, pubertal, post-pubertal), and (iii) age (within 12 months, where possible following ii). Based on the same exclusion criteria, the control group were recruited from online adverts on University of Bristol websites and on social media.

### Measures

#### Body composition

Height (to nearest 1mm) and weight (to nearest 1g) were measured using a stadiometer and calibrated weighing scales. The LMS Growth method was used to calculate body mass index standard deviation score (BMISDS; (40)). In addition, body composition was measured using multifrequency bioelectrical impedance (Tanita MC-780).

#### Resting metabolic rate (RMR)

RMR was measured using indirect calorimetry (Cosmed K4b2). The equipment was calibrated prior to each data collection to ensure accuracy. Once the appropriate-sized mask had been fitted, participants lay supine to relax and asked to stay quiet and still for the duration of the measurement (20 minutes). The flattest portion (10 minutes) of the collected data, as depicted graphically in the K4b2 software, was selected for the calculation of RMR in the Cosmed software.

#### Oral glucose tolerance test (OGTT) & Magnetic Resonance Imaging (MRI)

All participants consumed a glucose drink (oral glucose solution 1.75g/kg up to maximum of 75g). Computerised, subjective measures of hunger, fullness, thirst, and nausea were taken at baseline, 30-, 60-, 90- and 120- minutes post glucose consumption using visual analogue scales (100mm line with endpoints of ‘Not at all’ and ‘Extremely’). Blood samples were collected from patients only, via an intravenous cannula at baseline, 30-, 60-, 90- and 120-minutes post glucose consumption to measure glucose, insulin, ghrelin, GLP1 and PYY. Leptin, full blood count (FBC) and electrolytes (U&Es) were also measured at baseline.

Patients underwent two MRI scans in 30-minute scan blocks: at baseline prior to the glucose drink, and 60 minutes post glucose consumption, and compared with historical control data from a previous study with identical methodology (41). This research scan included a food cue reactivity task, which was given to the control participants outside of the MRI scanner. Note that the OGTT and associated methodology are reported here for transparency of the full study procedure; data will be presented elsewhere.

#### Hyperphagia questionnaire

The Hyperphagia Questionnaire (26) was used to measure hyperphagic eating behaviour across three subscales (drive, behaviour, and severity). Higher scores reflect a greater frequency and severity of food-related preoccupations and hyperphagic associated problems. To calculate a total hyperphagia score, the three subscales were summed. A modified, self-report version of the Hyperphagia Questionnaire was created for patients over 18 years old for this study.

#### Eating Behaviour questionnaire

The Child Eating Behaviour Questionnaire (CEBQ)(27) was used to measure appetitive traits associated with obesity risk in childhood. Participants over 18 years old completed the corresponding Adult Eating Behaviour Questionnaire (AEBQ) (42). This study also took an average of the three food approach traits and the four food avoidance traits to create a composite food approach score and a composite food avoidance score.

#### Quality of Life (QoL)

QoL was assessed using the Minneapolis-Manchester Quality of Life Survey of Health for Children and Young people (MMQL)(43). A parent of patients under 18 years completed the parallel parent version of the MMQL (44).

#### Ad libitum meal

The ad libitum lunch consisted of 14 vegetarian, hot and cold foods and two beverages served in excess and laid out in a standardised fashion. The total energy in the meal was 30,049KJ (37, 49, 11% of energy from fat, carbohydrate, and protein, respectively (Table 1). Each item was covertly weighed to the nearest gram by the investigator before and after the lunch. Patients remained in a room alone for 30 minutes and were asked to eat to appetite.

**Table 1.** Energy content and macronutrient composition of the ad libitum meal.

| Item | Amount (g) | Energy KJ | Fat | CHO | Protein |
| --- | --- | --- | --- | --- | --- |
| Macaroni cheese | 400 | 2733 | 29 | 66 | 30 |
| Wholemeal bread | 241 | 2388 | 8 | 87 | 17 |
| White bread | 208 | 2107 | 5 | 95 | 17 |
| Flora | 70 | 1170 | 32 | 0 | 0 |
| Cheese | 240 | 4140 | 84 | 1 | 61 |
| Humus | 115 | 1323 | 25 | 12 | 8 |
| red capsicum | 68 | 61 | 0 | 3 | 1 |
| Tomato | 127 | 77 | 0 | 4 | 1 |
| Fruit loaf | 500 | 6355 | 15 | 291 | 41 |
| OJ | 1000 | 1760 | 5 | 86 | 6 |
| Mandarin | 560 | 969 | 1 | 54 | 4 |
| Mayo | 38 | 1127 | 30 | 0 | 0 |
| Choc Chip cookies | 230 | 4743 | 55 | 143 | 13 |
| Walkers crisps | 50 | 1097 | 16 | 26 | 3 |
|  | Total | 30049 | 304 | 868 | 201 |
|  | %E |  | 37.4 | 49.1 | 11.4 |

#### Acceptability questionnaire(s)

Age-appropriate questionnaires (10 years old and under, 11-15 years old or 16 years and over) were designed specifically for this study to assess patient tolerability of the number and nature of study measures. Questions were worded to suit the participant’s age group and therefore not all questions were asked of all participants. Additionally, a parent/carer of the patient completed the parent/carer acceptability questionnaire. Questions were presented on a five-point rating scale; for patients ≤10 years and for those 11-15 years old, the five-point rating scale included different faces from a red sad face to a green happy face. The five-point rating scale for patients ≥16 years and parents ranged from 1 = not at all to 5 = extremely. Some questions required a yes or no response and included sections where the individual could comment freely.

### Procedure

Each participant was tested individually over the course of one morning at the Clinical Research Facility. The procedure, shown in Figure 1, for the control group was identical to that for patients with craniopharyngioma, except that the control group did not have an MRI scan or blood tests.

**Fig. 1.**
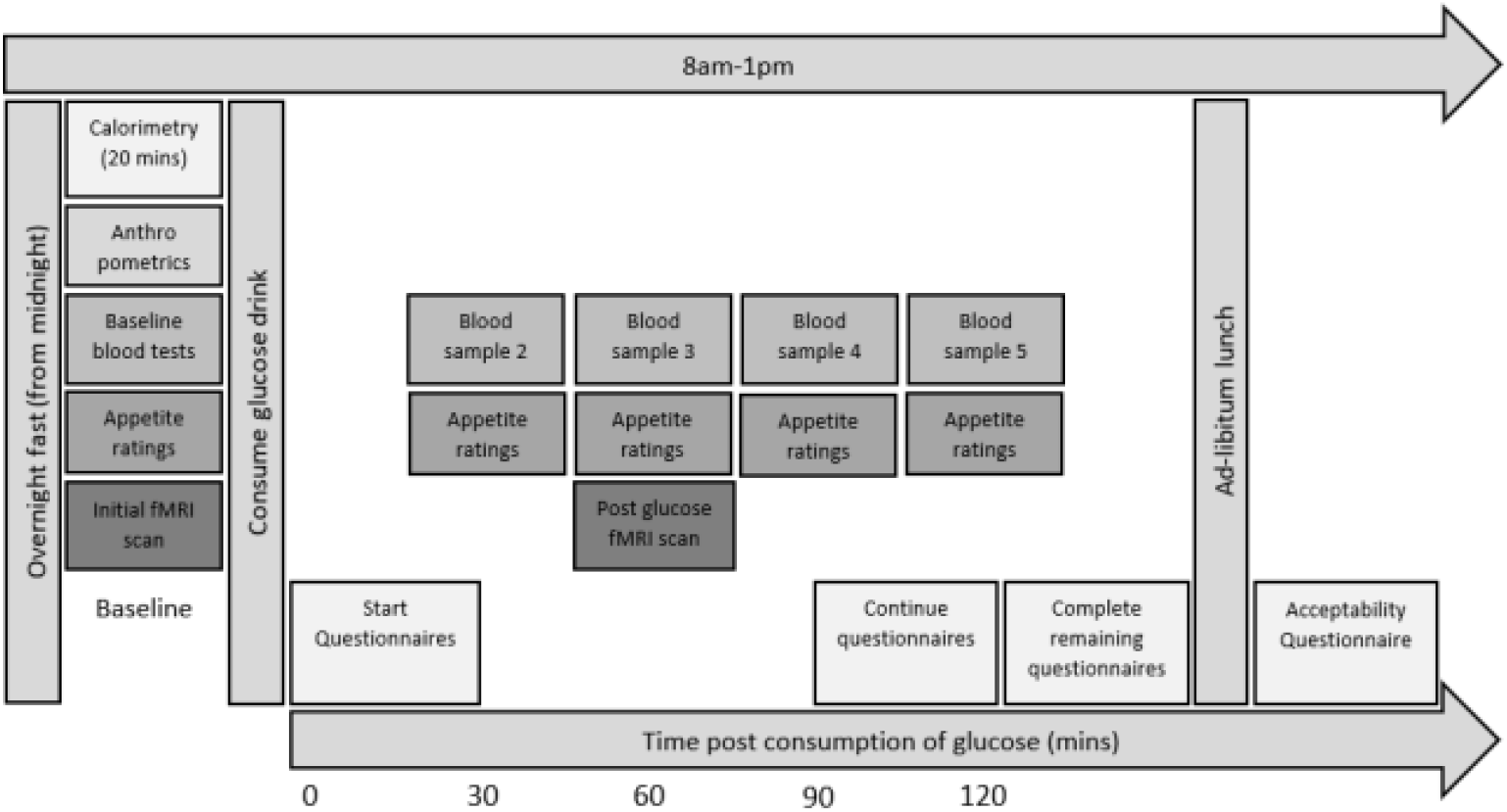
Study procedure

After an overnight fast of at least eight hours, participants received the above measures in the following order: body composition, resting metabolic rate, OGTT including MRI, appetite ratings and blood sampling), eating behaviour and QoL questionnaires, ad libitum lunch, and acceptability questionnaire. Participants were given a shopping voucher to thank them for their time and effort.

### Data processing and statistical analysis

The feasibility outcomes of the study were operationalised as follows: (i) patient data on key measures, including Paris grading of tumour, and pre- and post-op BMI; (ii) recruitment (no. of participants recruited and % of target sample); (iii) acceptability of the measures (scores of between 1 to 5 and comments) by participants and their parent/carer, and tolerance of the protocol (no. of participants who completed the full data set); (iv) preliminary data on the eating behaviour measures described above were summarized using medians (IQR) and frequencies where appropriate.

A priori, we decided to report effect sizes, due to concerns about null-hypothesis significance testing in small sample sizes and because p values do not indicate the magnitude of any reported effects [31–33]. Cliff’s delta was chosen as it has no assumptions on the underlying distribution and performs well for small sample sizes (n>10; (45)). Cliff’s delta and associated 95% confidence intervals were calculated using an available Excel macro (45). The macro notes that for the variables with a sample size of less than 10, the accurate estimation of the 95% confidence intervals is compromised. Descriptors of magnitude of effect size were 0.11 for small effect size, 0.28 for medium effect size and 0.43 for large effect size (45). For correlations, Kendall’s tau was used, and the following descriptors were applied for the magnitude of the correlation coefficient with r = 0.10 for small effect size, r = 0.30 for medium effect size and r = 0.50 for large effect size. Data was analyzed using SPSS version 24.

## Results

### Participant characteristics (objective 1)

Comprehensive searching of medical records, including the archives, allowed the full clinical characterisation of the 11 patients with craniopharyngioma to be presented in Table 2. Age at diagnosis ranged from 5 to 15 years, with surgical techniques used to treat the craniopharyngioma ranging from debulking, cyst aspiration and transsphenoidal resection. 9/11 patients had also received radiotherapy. Post-operatively, all patients had adrenocorticotropin hormone (ACTH) and growth hormone deficiencies, 10/11 had thyroid dysfunction and 6/11 had diabetes insipidus. There was wide variation in BMI SDS at 12 months post-op from underweight to obese categorisations, and the extent of hypothalamic damage varied across the Paris grading (Table 2).

**Table 2.** Clinical characterisation of the patients with craniopharyngioma.

| Participant no. | Sex | Age range at diagnosis (years) | Surgery | Radio-therapy received (yes/no) | Paris grading |  | BMI SDS |  | Diabetes insipidus |  | Thyroid dysfunction |  | ACTH deficiency |  | GH deficiency |  |
| --- | --- | --- | --- | --- | --- | --- | --- | --- | --- | --- | --- | --- | --- | --- | --- | --- |
|  |  |  |  |  | Pre-op | Post-op | Pre-op | 12m | Pre-op | 12m | Pre-op | 12m | Pre-op | 12m | Pre-op | 12m |
| 1 | F | 6-10 | Debulking | Yes | 2 | 1 | 3.20 | 3.4 | x | y | y | y | y | y | x | y |
| 2 | M | 6-10 | Debulking | Yes | 2 | 2 | 2.93 | 2.99 | x | x | x | y | x | y | x | y |
| 3 | M | 6-10 | VAD | No | 1 | 1 | -0.28 | -0.79 | x | x | y | y | y | y | y | y |
| 4 | M | 11-15 | Aspiration of cyst, VAD | Yes | 2 | 2 | -0.38 | -1.08 | x | x | x | y | y | y | x | y |
| 5 | F | 0-5 | Debulking | Yes | 2 | 2 | 1.43 | 3.43 | x | y | x | y | x | y | x | y |
| 6 | F | 6-10 | TSR | Yes |  | 0 | 0.09 | 0.94 | x | y | y | y | y | y | x | y |
| 7 | F | 11-15 | TSR | Yes | 0 | 0 | 0.17 | 0.72 | x | y | y | y | y | y | x | y |
| 8 | M | 11-15 | Aspiration of cyst, | Yes | 2 | 2 | 1.60 | 0.10 | x | x | x | x | x | y | x | y |
| 9 | M | 16-20 | TSR | No | 0 | 0 | -2.42 | -0.76 | y | y | y | y | y | y | y | y |
| 10 | F | 11-15 | Debulking, VAD | Yes | 1 | 2 | -0.84 | 0.61 | x | x | y | y | y | y | x | y |
| 11 | M | 6-10 | Aspiration of cyst | Yes | 2 | 2 | 0.18 | 1.34 | x | y | x | y | x | y | x | y |
| Overall<br>(n=11) | F:<br>5/11<br>M:<br>6/11 |  | Debulking: 4/11<br>Cyst aspiration:<br>4/11<br>TSR: 3/11 |  |  |  |  |  | 1<br>/11 | 6<br>/11 | 6<br>/11 | 10<br>/11 | 7<br>/11 | 11<br>/11 | 2<br>/11 | 11<br>/11 |
| Mean<br>(S.D) |  | 10.8<br>(3.49) |  |  |  |  | 0.52<br>(1.65) | 0.99<br>(1.66) |  |  |  |  |  |  |  |  |

S.D = standard deviation; F=female; M = male; VAD = ventricular access device; TSR = transphenoidal resection; Pre-op = Pre-operatively; Post-op = post- operatively; 12m = 12 months post-operatively; BMI SDS = body mass index standard deviation score; ACTH = adrenocorticotropin hormone; GH = growth hormone; y = yes, present.

### Recruitment (objective 2)

Eleven patients with craniopharyngioma were recruited over the period of two years from January 2019 until December 2020 (Figure 2). This time period included the first COVID-19 lockdown. Healthy-weight, matched control participants were recruited from October 2020 to October 2021. A target of 20 patients with craniopharyngioma was set and 55% of this target was reached during the study period with a marked reduction in the second year due to COVID-19 lockdown. 58% of those invited to take part did go on to participate (Figure 2). Reasons for not participating included patient illness, family health problems, and unwillingness to travel.

**Fig. 2.**
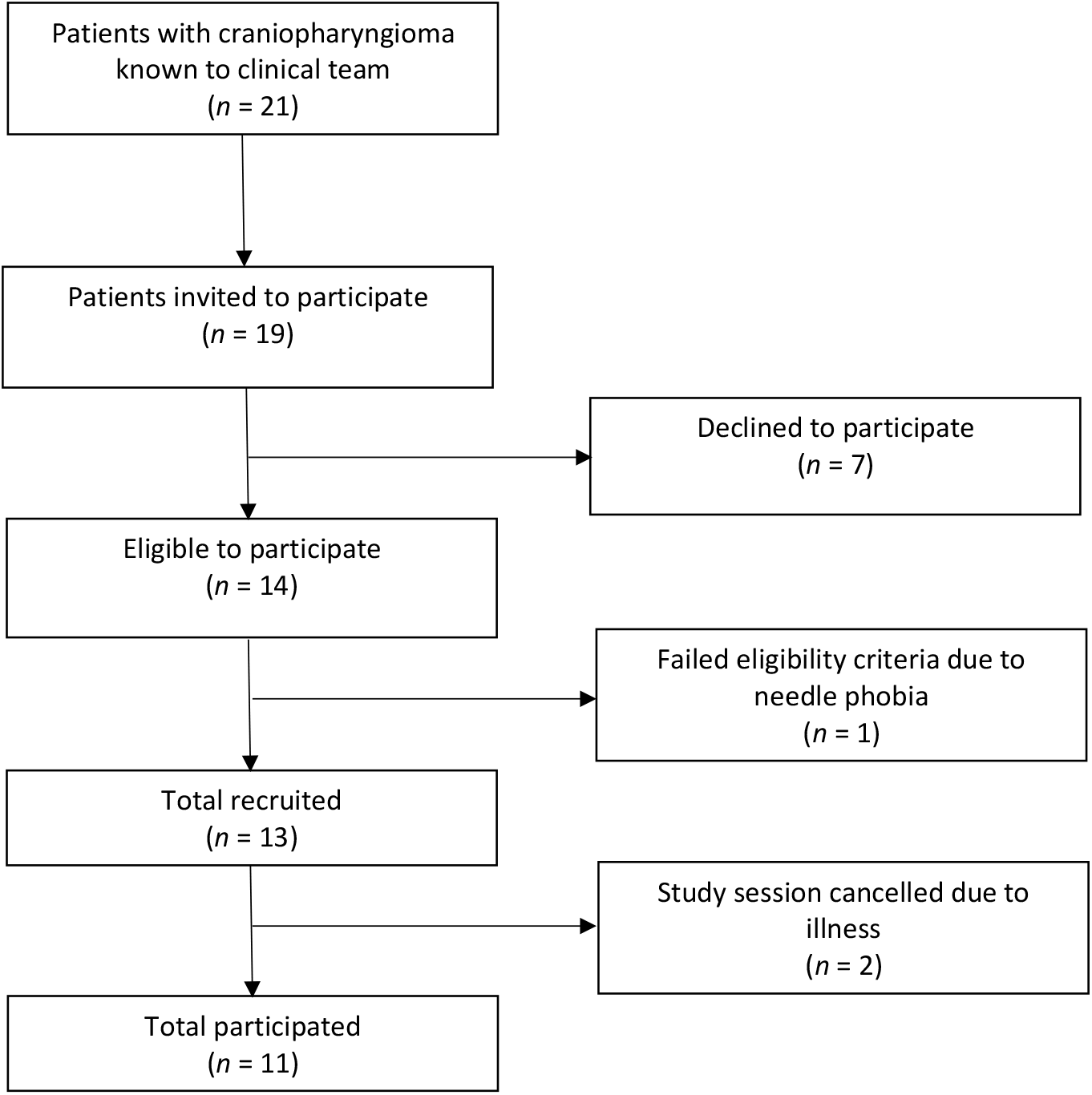
Participant flow chart

### Acceptability and tolerability of the measures (objective 3)

All patients (n = 11) and the matched controls (n = 11) completed the acceptability questionnaire suitable for their age (10 years and under n = 4, 11-15 years old n = 10, 16 years plus n = 8). A total of 5 parents (2 from patients and 3 from matched controls) of those tested over the age of 16 years were unavailable to complete the acceptability questionnaires. This resulted in a total of 17 parents (9 parents of patients and 8 parents of the matched controls) who completed the acceptability questionnaires.

Overall, participants found the various aspects of the study highly acceptable and well tolerated. Table 3 reports missing data, acceptability, and tolerability of each measure. The number of patients and controls who completed each of the measures (between 80-100%) is reported in Table 4.

**Table 3.** Feasibility and acceptability of included measures.

| Measure | Data issues | Acceptability rating (median, 95% C.I.) | Patient or Parent comments / notes (age range in brackets) |
| --- | --- | --- | --- |
| Body composition | n=4 missing for the trunk measurement due to data recording issues. | Patient = 5, 95% CI: 4-5; Control = 5, 95% CI: 4-5 | *"I also found it interesting to learn about the children's muscle mass"* (parent of 11-15y, M) |
| Resting metabolic rate | n=3 not possible to collect data due to technical issues with the equipment. Some found lying still for 20 minutes to collect the data difficult. | Patient = 5, 95% CI: 4-5; Control = 4.5, 95% CI: 3-5 | *"it was wierd but it was fine"* (11-15yr, F) |
| Oral glucose tolerance test | n=1 not possible to cannulate | Glucose drink: Patient = 3, 95% CI: 2-5; Control = 4, 95% CI: 2-4<br>Bloods: Patient = 4, 95% CI: 4-5 | *"it was gloopy, too sweet and not nice"* (11-15yr, F)<br>*"Didn't taste very nice"* (11-15yr, M)<br>*"obviously I didn't want to do it, but it didn't hurt so it was fine"* (11-15yr, F)<br>*"it will hopefully negate the need for my next blood test which is a bonus!"* (21-25yr, F) |
| MRI | n=2 data not collected due to bioplate and suspected surgical clips | Bloods: Patient = 4, 95% CI: 3-5 | *"the first one was uncomfy but the other one was fine"* (11-15yr, F) |
| Questionnaires | No issues | Patient = 5, 95% CI: 4-5; Control = 4, 95% CI: 4-5 | If the participant was 18 years or older, they were not required to have a parent or carer present in order to complete the acceptability or QoL parent/carers data |
| Ad libitum lunch | n=1 control participant reported food allergies after testing had started so their data were excluded as it was incomplete | Patient = 5, 95% CI: 4-5; Control = 5, 95% CI: 4-5 | *"more choice"* (11-15yr, M)<br>*"There was a wide range however the eating environment felt artificial to me"* (21-25yr, M)<br>*"I really liked the oranges"* (11-15yr, M) |
|  |  |  | *"Great food" (11-15yr, M)* |
| How **interesting** did you find the project overall? | No issues | Patient = 4, 95% CI: 3-5; Control = 4, 95% CI: 4-5 | *"I have always been intrigued around my eating habits since my op so it has really interested me being asked to do this research project" (16-20yr, F)* |
| Fasting overnight | No issues | Patient = 4, 95% CI: 3-5; Control = 4, 95% CI: 3-5 | *"I got very hungry and it was hard to have a different routine" (11-15yr, F)*<br>*"anticipation worse than actually missing it\*" (11-154yr, M) \*breakfast* |
| Travelling and staying overnight | No issues | Patient = 3, 95% CI: 3-5; patients only. |  |
| How important do you think it is to investigate how eating behaviour in childhood craniopharyngioma? | No issues | Free text comments only | *"it is easy to focus on the endocrine/surgical side of things without investigating how QoL can be improved. Eating behaviour is something which can have a wide impact, not only on someone's health but also on their mental and social wellbeing hence the need for such studies" (21-25yr, F)*<br>*"Very important. We're struggling with participant's diet and appetite." (parent of 6-10yr, F)*<br>*"It is clearly very important, there is a link between craniopharyngioma and obesity, yet it appears no real explanation as to why this is." (parent of 6-10yr, M)* |
| Reported changes in eating | No issues | Free text comments only | *"... eating habits have definitely changed since his diagnosis. He clearly talks more about food and always wants to know what's for tea!" (parent of 6-10yr, M)* |
| Benefits of taking part | No issues | Free text comments only | *"the care ... has received throughout her childhood and adult life will have benefitted from research projects in the past. To be involved in further research will hopefully help future* *sufferers of craniopharyngioma.”*<br>(parent of 21-25yr, F). |

**Table 4.** Comparison of the craniopharyngioma patient and control groups.

| Measure | Patients with craniopharyngioma |  | Matched-control participants |  | Effect size |
| --- | --- | --- | --- | --- | --- |
|  | Median (IQR) or frequency | N | median/IQR or frequency | N | Cliff's delta (95% C.I.s) |
| ***Demographic data*** |  |  |  |  |  |
| Sex (M/F) | 6/5 | 11 | 6/5 | 11 | na |
| Age (years) | 14.0 (9.0) | 11 | 12.0 (10.0) | 11 | 0.1 (0.5, -0.4) |
| Pubertal stage (pre/ during/post) | 5 / 2 / 4 | 11 | 5 / 2 / 4 | 11 | na |
| Ethnicity | 1 Pakistani; 10 British | 11 | 1 Pakistani; 1 Asian, 9 British | 11 | na |
| ***Body composition*** |  |  |  |  |  |
| Height SDS | 0.53 (3.1) | 11 | 0.2 (1.8) | 11 | 0.8 (0.5, -0.4) |
| Weight SDS | 1.9 (3.0) | 11 | -0.2 (1.2) | 11 | 0.5 (0.8, -0.1) |
| BMI SDS | 1.4 (3.0) | 11 | -0.5 (1.3) | 11 | 0.6 (0.8, 0.0) |
| Fat mass (%) | 29.9 (23.6) | 10 | 18.2 (7.2) | 11 | 0.5 (0.8, -0.1) |
| Fat mass (kg) | 16.0 (21.1) | 10 | 6.7 (1.3) | 11 | 0.5 (0.8, 0.0) |
| ***Energy expenditure*** |  |  |  |  |  |
| Resting metabolic rate (RMR) | 1874.0 (839.0) | 10 | 1791.0 (382.0) | 9 | 0.1 (0.6,-0.4) |
| ***Energy intake (ad libitum meal)*** |  |  |  |  |  |
| Energy (kJ) | 4886.0 (3426.0) | 11 | 4783.0 (2193.0) | 10 | 0.0 (0.5, -0.5) |
| Fat (g) | 50.0 (30.0) | 11 | 50.0 (21.8) | 10 | 0.0 (0.4, -0.5) |
| Fat (% energy) | 40.0 (11.0) | 11 | 38.0 (9.5) | 10 | 0.0 (0.5, -0.5) |
| CHO (g) | 148.0 (79.0) | 11 | 129.5 (71.5) | 10 | 0.1 (0.5, -0.4) |
| CHO (% energy) | 50.0 (8.0) | 11 | 45.5 (10.5) | 10 | 0.2 (0.6, -0.3) |
| Protein (g) | 28.0 (27.0) | 11 | 37.0 (13.5) | 10 | -0.2 (0.3, -0.6) |
| Protein (% energy) | 9.0 (4.0) | 11 | 11.5 (3.8) | 10 | -0.3 (0.2, -0.7) |
| ***Hyperphagia questionnaire*** |  |  |  |  |  |
| Hyperphagia: Total | 23.0 (15.0) | 11 | 16.0 (8.0) | 11 | 0.5 (0.8, 0.0) |
| Hyperphagia: behaviour | 10.0 (9.0) | 11 | 6.0 (3.0) | 11 | 0.5 (0.8, 0.0) |
| Hyperphagia: drive | 9.0 (6.0) | 11 | 7.0 (4.0) | 11 | 0.4 (0.8, -0.1) |
| Hyperphagia: severity | 4.0 (2.0) | 11 | 2.0 (1.0) | 11 | 0.7 (0.9, 0.2) |
| ***Adult/Child Eating Behaviour Questionnaire*** |  |  |  |  |  |
| Enjoyment of Food | 4.7 (1.3) | 11 | 4.0 (1.8) | 11 | 0.3 (0.7, -0.2) |
| Emotional overeating | 2.3 (1.5) | 11 | 2.0 (0.9) | 11 | 0.3 (0.7, -0.2) |
| Food responsiveness | 3.5 (2.0) | 11 | 2.5 (0.7) | 11 | 0.3 (0.7, -0.3) |
| Slowness in eating | 2.5 (2.5) | 11 | 2.8 (1.8) | 11 | -0.2 (0.3, -0.6) |
| Food approach | 3.4 (1.6) | 11 | 2.7 (0.9) | 11 | 0.4 (0.7, -0.1) |
| Satiety responsiveness | 2.5 (1.4) | 11 | 3.3 (1.6) | 11 | -0.4 (0.1, -0.8) |
| Emotional undereating | 2.5 (2.1) | 11 | 3.0 (1.1) | 11 | -0.1 (0.4, -0.5) |
| Food fussiness | 2.7 (2.3) | 11 | 2.8 (1.7) | 11 | -0.2 (0.3, -0.6) |
| Food avoidance | 2.7 (0.8) | 11 | 3.0 (0.5) | 11 | -0.4 (0.1, -0.7) |
| ***Quality of Life (MMQL)*** |  |  |  |  |  |
| Participant total | 65.5 (28.5) | 11 | 78.5 (20.7) | 11 | -0.3 (0.2, -0.7) |
| Participant physical | 80.0 (35.0) | 11 | 95.0 (25.0) | 11 | -0.5 (0.0, -0.8) |
| Participant emotional | 60.7 (32.1) | 11 | 71.4 (10.7) | 11 | -0.2 (0.3, -0.6) |
| Participant social | 91.7 (20.8) | 11 | 83.3 (12.5) | 11 | 0.1 (0.6, -0.4) |
| Participant school | 57.1 (39.3) | 11 | 75.0 (35.7) | 11 | -0.3 (0.2, -0.7) |
| Participant physical appearance | 50.0 (50.0) | 11 | 81.3 (56.3) | 11 | -0.2 (0.3, -0.6) |
| Parent total | 59.5 (13.8) | 9 | 83.6 (18.9) | 8 | -0.9 (-0.5, -1.0) |
| Parent physical | 45.0 (28.0) | 9 | 92.5 (20.0) | 8 | -0.8 (-0.2, -1.0) |
| Parent emotional | 64.3 (28.6) | 9 | 64.3 (8.9) | 8 | -0.1 (0.5, -0.6) |
| Parent social | 79.2 (35.4) | 9 | 91.7 (14.6) | 8 | -0.5 (0.0, -0.8) |
| Parent school | 46.4 (57.2) | 9 | 92.9 (18.8) | 8 | -0.8 (-0.2, -0.9) |
| Parent physical appearance | 37.5 (59.4) | 9 | 75.0 (34.4) | 8 | -0.3 (0.3, -0.7) |

### Preliminary data with matched controls (objective 4)

As intended, the groups were well-matched on sex, pubertal stage, and age (Table 4).

### Body composition, energy intake and energy expenditure

Patients had a higher BMI SDS, and absolute and percentage fat mass compared to controls with a large effect size for each measure. No difference was found between groups in RMR (Table 4). Total energy intake at the *ad libitum test* meal did not differ between groups. When macronutrient intake was expressed as a percentage of total energy, patients consumed more carbohydrate and fat, and less protein than controls.

### Eating behaviour questionnaires

There was greater hyperphagia in the patients compared to controls, with the largest effect size for the hyperphagia severity subscale (d=0.7; Table 4). More time spent talking or engaging with food and a greater extent to which food interfered with daily functioning was reported in patients. Results from the A/CEBQ showed greater food approach behaviours (such as enjoyment and responsiveness to food and emotional overeating) in patients with a medium to large effect size (Table 4). By contrast, greater food avoidance behaviours, such as slowness in eating, satiety responsiveness and food fussiness, were reported for controls, with medium to large effect sizes (Table 4).

### Quality of life (QoL)

QoL of participants overall was lower in patients than controls, both as reported by the young people with a medium effect size, and by their parent/carer with a large effect size (Table 4). Lower QoL was reported by parents of patients compared to their children (d = 0.26), whereas young people in the control group rated their QoL lower than their parents (d = -0.22). Patients reported that their health restricted their activities (‘physical functioning’ subscale) to a greater extent than controls, whereas the largest differences in parents/carer ratings between groups were found for the physical and social functioning (having close friends with similar interests) and functioning at school (concentration and difficulty with schoolwork) subscales, with lower QoL on these subscales for the patients.

### Relationship between eating behaviour measures and BMI-SDS

A positive correlation between food approach behaviours and BMISDS was found with a large effect size in patients but not matched controls (Table 5). A positive correlation was found between hyperphagia total score and BMISDS in patients, with a much weaker relationship in controls. Conversely, a negative correlation of medium effect size was seen between food avoidance behaviours and BMISDS in patients, with only a weak relationship in controls.

**Table 5.** Relationship (Kendalls tau) between eating behaviour measures and BMI-SDS.

|  | COSMED full dataset RMR (kcal/day) | Energy intake (kJ) | Food Approach | Food Avoidance | Hyperphagia total score |
| --- | --- | --- | --- | --- | --- |
| Craniopharyngioma group | -0.02 | 0.27 | .55 | -0.38 | 0.43 |
| Matched controls | 0.25 | 0.41 | 0.04 | 0.11 | 0.23 |

### Relationship between QoL, eating behaviour and BMISDS

Only weak evidence was found for negative correlations between participant-reported QoL total score and BMISDS (patients = -0.20; controls = 0.13, small effect sizes) and parent-reported QoL total score and participants BMISDS (patients = -0.25, medium effect size; controls = -0.18, small effect size). In patients, there was little evidence for a relationship between their self-reported QoL scores and energy intake or expenditure or eating behaviour measures (small effect sizes only) (Table 6). For parent/carer QoL measure, relationships of medium effect size were found with food avoidance and hyperphagia scores, whereby greater food avoidance behaviours were positively correlated, and hyperphagia scores negatively correlated, with QoL.

**Table 6.** Relationship (Kendalls tau) between eating behaviour measures and QoL.

|  |  | COSMED full dataset RMR (kcal/day) | Energy intake (kJ) | Food Approach | Food Avoidance | Hyperphagia total score |
| --- | --- | --- | --- | --- | --- | --- |
| Patient group | Participant reported QoL (n=11) | -.18 (n=10) | 0.09 | -0.08 | 0.17 | -0.11 |
|  | Parent/carer reported QoL (n=9) | -0.07 (n=8) | -0.09 | -0.09 | 0.42 | -0.40 |
| Matched controls | Participant reported QoL (n=11) | 0.37 (n=9) | 0.00 (n=10) | -0.34 | 0.15 | -0.39 |
|  | Parent/carer reported QoL (n=8) | -0.20 (n=6) | -0.26 | -0.26 | 0.04 | -0.39 |

## Discussion

This study has demonstrated it is feasible and acceptable to conduct multi-component, eating behaviour research in young people with craniopharyngioma. While recruitment of adequate patient numbers from two sites was challenging, 58% of those invited participated. Overall, the included measures were deemed highly tolerable by participants and their parents and have provided preliminary data to begin to investigate in one cohort, the consequences of hypothalamic damage on energy intake, energy expenditure and eating behaviour, and the impact these may have on quality of life.

### Feasibility of research into eating behaviour and obesity in young people with craniopharyngioma

Previous research has indicated that recruitment can be challenging in studies of craniopharyngioma due to the complexity of the condition (46). Indeed, considering the heterogeneity of disease and treatment complications following craniopharyngioma (47), it is unsurprising that 10 of the 21 patients initially identified did not align with the study criteria. This included those with severe visual problems who could not be included due to the requirements of completing the task in the MRI scanner. Other reasons for not participating were given by invited families, such as difficulty of scheduling clinical sessions at the same time as the research session, and anxiety surrounding having blood taken were noted. Moreover, the COVID-19 pandemic suspended testing from March-October 2020. Future studies should recruit from several specialist centres to obtain sufficient patient numbers and should allow access to participate in different aspects of the study if not all criteria are met.

Participants broadly found the protocol both tolerable and acceptable. Aspects of the OGTT, including the glucose drink, blood tests, and missing breakfast to be fasted were the least tolerable components of the protocol. The meal elicited the most comments with several participants reporting they enjoyed it, others asking for more choice, with another commenting that the environment felt artificial. The experience of taking part was valued by young people and their parents alike, with many commenting on the importance, yet lack of understanding, of eating behaviour for those with craniopharyngioma. Several parents reported changes and difficulties with their child’s eating and could also see the wider benefits of taking part.

### Preliminary data comparing those with and without craniopharyngioma

Analysis of the preliminary data was conducted to inform a larger, multi-centre trial to assess interventions designed to improve eating behaviour in young people with hypothalamic obesity. To the authors’ knowledge, only one study has conducted an *ad libitum* meal to measure dietary intake in craniopharyngioma patients (17). However, in the current small, feasibility study, only a weak relationship between total energy intake and BMISDS was found in patients with craniopharyngioma, suggesting that it is difficult to assess whether the amount of food consumed is a primary driver of obesity for these patients using an acute measure from a standard meal alone.

By contrast, questionnaire measures of eating behaviour consider habitual eating traits over a longer period (27, 42). Preliminary evidence was found for greater food approach behaviours, including hyperphagic behaviours of greater severity (26), in patients with craniopharyngioma compared to sex, pubertal stage and age matched controls. Shoemaker and colleagues (17) also found similar levels of hyperphagia and variability in scores across patients using the Hyperphagia questionnaire, with high levels of enjoyment of food, like the current study. Together, these findings suggests that young people with craniopharyngioma experience enjoyment from their food, but in some cases, this heightened food responsivity could lead to problematic behaviours, such as stealing food or anger outbursts if access to food is removed. Moreover, both food approach and hyperphagia were positively correlated with BMISDS with a medium to strong effect size, suggesting that the greater drive patients with craniopharyngioma have to engage in food approach and consumption behaviours, the higher level of BMISDS. Moderating hyperphagia scores, therefore, could be an informative primary outcome for a future trial. A power calculation (using G*Power, assuming 80% power to detect a difference between groups) based on the effect size from this study (d=1.0) suggests that a minimum of 27 participants would be required in each group. These numbers, together with the high patient acceptability of the questionnaire measures, suggests hyperphagia score change (26), could be a feasible outcome for a future trial which would allow the exploration of other confounding factors, such as hypothalamic damage and history of surgery and radiotherapy.

For there to be a clear patient benefit, it is also important to assess the impact on QoL. Interestingly, QoL as rated by the parents of young people in this study was found to be related to these same hyperphagic eating behaviours with a medium effect size; that is, QoL was rated as lower for those young people with greater hyperphagic and food approach tendencies. Moreover, QoL was rated higher by parents of those who engaged in more food avoidance behaviours. Yet, this was not the case for QoL as rated by the young people themselves. One could speculate, that as they experience enjoyment from food, they do not always make the link between their eating and QoL, especially as in this sample, there was only a weak relationship between BMISDS and QoL. This is an interesting concept to explore further in future research.

When considering which other measures to include in a future trial, the findings from this study suggest that the body composition measures show clear, informative differences between patients and controls and were broadly feasible and acceptable to participants. Measurement of energy expenditure was more challenging in this young patient group. While no difference in resting metabolic rate was found between groups, contrary to previous studies (29-31), discrepancies between our findings and the previous work could be due to differing levels of BMISDS or hypothalamic damage and a smaller sample size in our feasibility study or the characteristics of the comparison groups (healthy weight or obese). This point highlights the inconsistencies of research into eating behaviour and the development of obesity within this patient group, both in terms of study design and findings to date, and provides the impetus to design larger, multi-centre trials with appropriate control groups within international networks of collaborators to further this field. The MRI and OGTT measures were also well tolerated, and while challenging to collect complete datasets, should be considered for future research designed to investigate the neural and physiological mechanisms underlying the development of obesity, and could be applied to further understand mechanisms of action of new drug therapies, such as GLP1 agonists, in this patient group (17). Full characterization of the patients is important in any future trial to ascertain whether behavioural and pharmacological interventions are differentially effective based on key factors, such as hypothalamic damage, development of obesity post-surgery and endocrine abnormalities.

### Limitations

There are missing data from several of the included measures; specifically, there were some user-related problems with the energy expenditure and body composition equipment, and difficulties in the cannulation of one of the young patients with obesity. However, this feasibility study has allowed the identification and response to potential difficulties of conducting this type of in-depth clinical study to take forward. It is important to note when considering the above findings that the control group reported in this paper was purposefully not matched on BMISDS for this study. This design choice has precedence in the literature (e.g. 30). The original design of the study was to recruit two control participants for each patient, one of healthy weight and one in the obese category; however, it proved very challenging to recruit young people with obesity to take part during the period of this research project, which coincided with the COVID-19 pandemic. All those who volunteered and matched a patient on sex, pubertal stage, and age, were of a healthy weight. However, the larger study, including MRI and OGTT measures, has a historical control group, comprising young people living with and without obesity, who took part in a previous MRI study investigating food-cue reactivity around a OGTT (41). The overlapping measures from these datasets, including OGTT results and food-cue reactivity during functional neuroimaging, will be presented in a future paper.

### Conclusion

The findings from this study suggest that the food approach behaviours, including food responsiveness and enjoyment of food, and for some patients, more severe instances of hyperphagic behaviours, show the strongest link with BMISDS. Future research should investigate how these specific behaviours may change over time, from pre to post surgery, depending on factors such as hypothalamic damage. Food approach behaviours may be useful targets for interventions to help patients with their eating; food-related, response inhibition training for example, has been shown to reduce consumption of high energy dense foods in adults and children (48, 49). Conversely, encouraging food avoidance behaviours, such as slowness in eating or responsiveness to satiety, may also serve to reduce food intake and improve health-related quality of life. It is important for future research, therefore, to ensure that the design of the study does allow the key questions to be addressed and does not exclude the participants with relevant symptoms. A key strength of this work is including the patient’s and their parent’s voice early in the research pathway; this area of research is important to patient and their families, and they are strongly motivated to support research which aligns closely with their key concerns.

## Data Availability

All data generated or analyzed during this study are included in this article. Further enquiries can be directed to the corresponding author.

## Statements

## Acknowledgement

We thank the young people and parents for participating in this study. We also thank Aileen Wilson and the staff at CRICBristol/Clinical Research Facility.

## Statement of Ethics

### Study approval statement

This study protocol was reviewed and approved by the South West - Frenchay Research Ethics Committee, approval number 18/SW/0235 (Nov 2018).

### Consent to participate statement

In accordance with HRA and General Medical Council guidance, written informed consent was obtained from participants if aged 16 years or over, or by their parent if 15 years or younger. Participants 15 years or younger provided assent to participate. This consent process is detailed in the methods section and was approved by the above Research Ethics Committee.

## Author Contributions

EH, FL, LC, KN, and JHS designed the study; EH, KN, FL, RLE, KH, SS, HB, NN, RE, TTM collected the data; EH, FL, RLE, SS, HB, NN analysed the data; EH wrote the first draft of the paper; all authors commented and contributed to the final manuscript.

